# Overcoming Emmetropic Presbyopia by Optimal Keratoplasty (Opti-K)

**DOI:** 10.1101/2025.08.16.25333839

**Authors:** K. Jonathan Rodgers, Harry G. Glen, Michael J. Berry

## Abstract

**PURPOSE:** To evaluate the safety and efficacy of the VIS Optimal Keratoplasty (Opti-K) device and procedure for treating eyes with emmetropic presbyopia to achieve uncorrected near visual acuity (UNVA) improvement.

**SETTING:** Bahamas Vision Centre, Nassau, New Providence, The Bahamas

**METHODS:** 145 eyes of 75 patients with emmetropic presbyopia (MRSE = -0.25 to +0.75 D; mean add and standard deviation (SD) = 1.89 (0.35 D) received primary treatments (Txs) by the VIS NTK Optimal Keratoplasty (Opti-K) device; 66 and 16 eyes also received secondary and ternary Txs, respectively. Patients were treated to produce best uncorrected near visual acuity (UNVA) in both eyes using a multifocal vision protocol. Safety and efficacy measures with follow-up (f/u) extending to 24 months post-Tx were analyzed.

**RESULTS:** <u>Safety</u> - There were no clinically significant adverse events or complications. Changes in intraocular pressure (IOP) and astigmatism (A) were not statistically significant (*p* < 0.05). Increase of central corneal thickness (CCT) was not clinically significant.

<u>Efficacy</u> - Geometric mean (gm) (SD) binocular UNVA improvements corresponded to 4.0 (2.0) lines gained at 1day′ post-secondary Tx (*p* < 10^-6^), with gradual reduction of improvement to 1.8 (1.5) lines gained at 24m′ (*p* = 0.002). Gm UDVA was conserved or even improved at most f/u times. Patient neuroadaptation to multifocality occurred immediately post-Tx. Mean durations (SD) of functional (>20/40) binocular UNVA outcomes were 10.8 (9.1) m for primary Txs only and 15.2 (10.3)m for primary plus secondary Txs.

**CONCLUSIONS:** In eyes with emmetropic presbyopia, optimal keratoplasty (Opti-K) is safe and efficacious for improving UNVA while retaining (or improving) UDVA. The procedure is noninvasive, simple, rapid, comfortable and repeatable. Although UNVA improvement is temporary, additional Opti-K Txs can be repeated whenever needed to maintain functional uncorrected near visual acuity.

**PRÉCIS:** In a retrospective observational study, Optimal Keratoplasty (Opti-K), a noninvasive procedure for vision improvement, provided safe and efficacious multifocal vision to emmetropic presbyopes, who gained uncorrected near visual acuity while maintaining (or improving) uncorrected distance visual acuity.

## INTRODUCTION

Presbyopia (aka age-related focus dysfunction) is a leading cause of vision impairment worldwide, adversely affecting over a billion people [Ref. 1]. Presbyopia is a progressive condition, typically starting at *ca*. age 40, and leading to ever increasing loss of uncorrected near visual acuity (UNVA) until lens flexibility is completely lost [Ref. 2].

Numerous surgical and pharmacological approaches have been developed for the treatment of presbyopia [Ref. 3]. All of these approaches are limited in efficacy and in some cases have safety problems, but VIS Optimal Keratoplasty (Opti-K) treatment has a very safe and efficacious clinical profile [Ref. 4]. The VIS Opti-K device and procedure are being evaluated in an U.S. FDA Clinical Trial, currently starting Phase 3 (pivotal, definitive) study; through Phase 2, treatment outcomes for only 30 eyes with emmetropic presbyopia have been obtained [Ref. 5]. Opti-K treatment outcomes for a small number of presbyopic patients in pilot studies have also been obtained [Refs. 6 and 7].

The purpose of this study was to evaluate the safety and efficacy of the VIS Optimal Keratoplasty (Opti-K) device and procedure to overcome emmetropic presbyopia by providing UNVA improvement.

## PATIENTS and METHODS

This retrospective observational cohort study (registered as NCT06702020 at https://ClinicalTrials.gov) was completed in conformance with ethical principles of the World Medical Association Declaration of Helsinki. The study protocol was approved by the Ethics Committee of the Bahamas Ministry of Health. Opti-K treatment (Tx) was provided to 145 eyes of 75 patients (55**♀**, 20**♂**; mean age: 50.0 ± 5.4 y, range 40 to 68 y) with emmetropic presbyopia (EP). Inclusion criteria included age ≥ 40 y; manifest refraction, spherical equivalent (MRSE) = -0.25 to +0.75 diopter (D) (mean: +0.28 ± 0.27 D) with ≤ 0.75 D cylinder; and corrected distance visual acuity (CDVA) ≥ 20/30. Exclusion criteria included nystagmus; uncontrolled uveitis, severe blepharitis, lagophthalmos; moderate to severe dry eye disease; corneal pathology and/or shape disorders such as keratoconus; previous ocular surgery; autoimmune disease; immunocompromised status; chronic allergic reactions; glaucoma; and diabetes. Pre-Tx, all eyes had presbyopia with mean add of +1.89 ± 0.35 D (range: +0.75 to +2.5 D) and with corrected near visual acuity (CNVA) ≥ J1 (0.1 logMAR, 0.8 decimal, 20/25 Snellen).

Examinations were performed pre-Tx and at post-Tx times (1d, 1w, 1m, 3m, 6m, 12m, and 24m) for both primary and non-primary Txs. Examination measurements included uncorrected (U) and corrected (C) distance and near visual acuities (UDVA, UNVA, CDVA, CNVA, binocular UDVA, binocular UNVA) - all measured using projected letter charts for distance VA and a Rosenbaum chart for near VA (Jaeger values were converted to logMAR, decimal and Snellen values for reporting); subjective manifest refraction; corneal topography (using an Atlas Model 991 corneal topographer; Zeiss Humphrey Systems, Dublin, CA); intraocular pressure (by Goldmann applanation tonometry); pachymetry (using Model DGH-500 ultrasonic pachymeter; DGH Technology, Eaton, PA); slit-lamp biomicroscopic evaluation (including Na fluorescein staining to visualize epithelial damage); subjective discomfort assessment; and subjective satisfaction with binocular UDVA/UNVA.

All eyes were treated with a prototype VIS Optimal Keratoplasty (Opti-K) device [Ref. 6]. The device included a continuous wave (cw) laser source operating at 1.93 µm wavelength, an optical delivery system comprising 16 optical fibers in a handpiece with a fixed Tx pattern, and a sapphire applanation window/suction ring (SAWSR) that is mounted on the cornea. Figure 1 shows the primary Tx pattern which consisted of two rings (6.0 and 7.2 mm centerline diameters; in some cases, two rings of 5.5 and 6.7 mm or 5.0 and 6.2 mm centerline diameters were used – there was no significant difference in outcomes for the three sets of Tx parameters so only combined data are reported) of Tx spots (0.5 mm diameter) arranged in symmetrical octagons with spots on radials; for secondary Tx, the Tx pattern was rotated *ca*. 22° from the primary Tx orientation shown in Figure 1. For ternary and quaternary Txs, the Tx pattern was rotated back to primary and secondary positions, respectively. The same Tx patterns were used for 66, 16 and 4 eyes receiving secondary, ternary and quaternary Txs, respectively. Two eyes of one patient received 10 Txs over a five-year period. Figure 2 shows a treated eye immediately post-primaryTx; the opacifications in treated spots fade and are not cosmetically significant.

**Figure 1.**
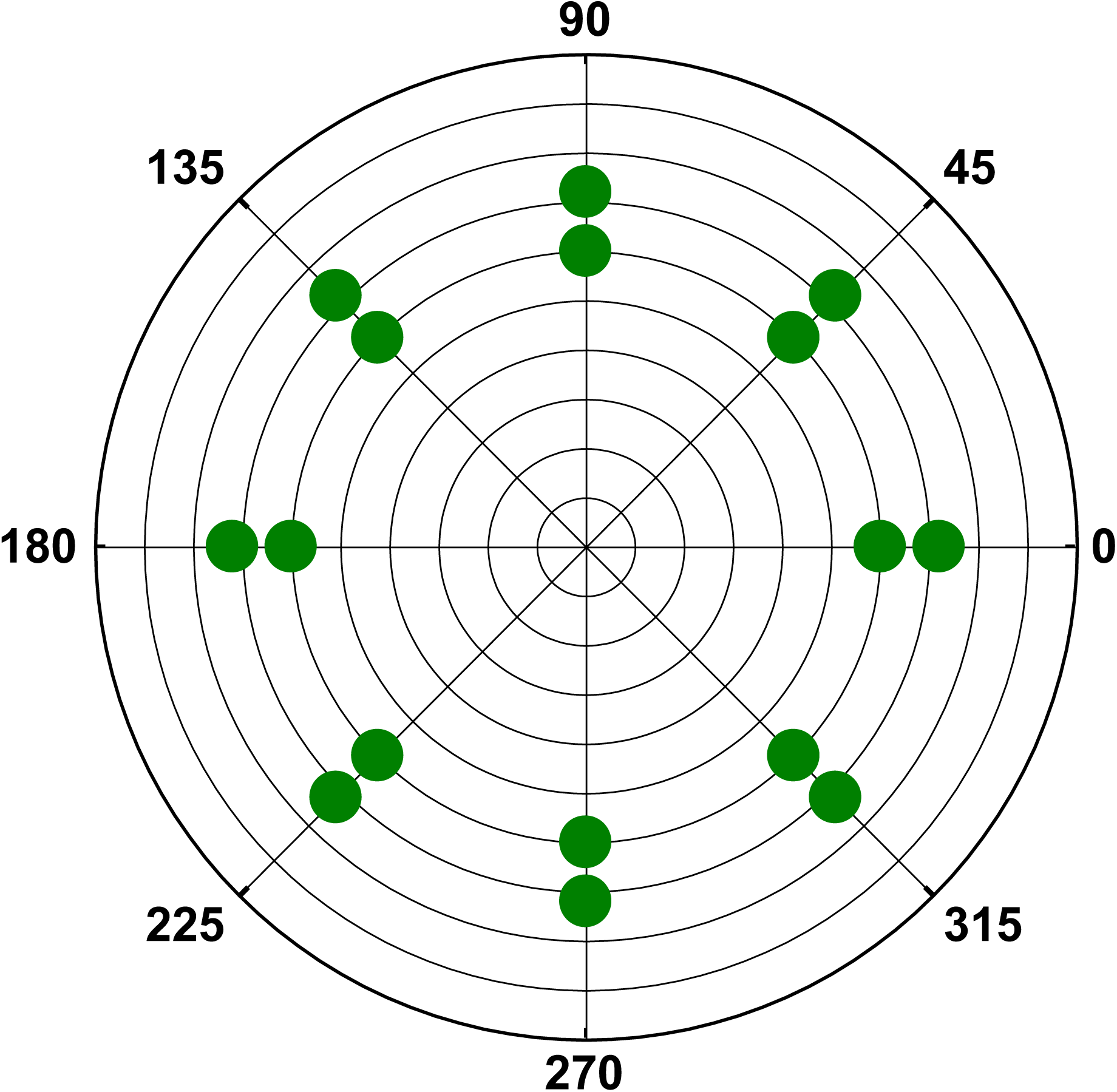
- 16-Spot eight-fold symmetry optimal keratoplasty (Opti-K) primary treatment pattern. Concentric rings are at 1 mm diameter intervals and are centered with respect to the pupillary center; treatment spots are located symmetrically and radially on rings at 6.0 and 7.2 mm centerline diameters.

**Figure 2.**
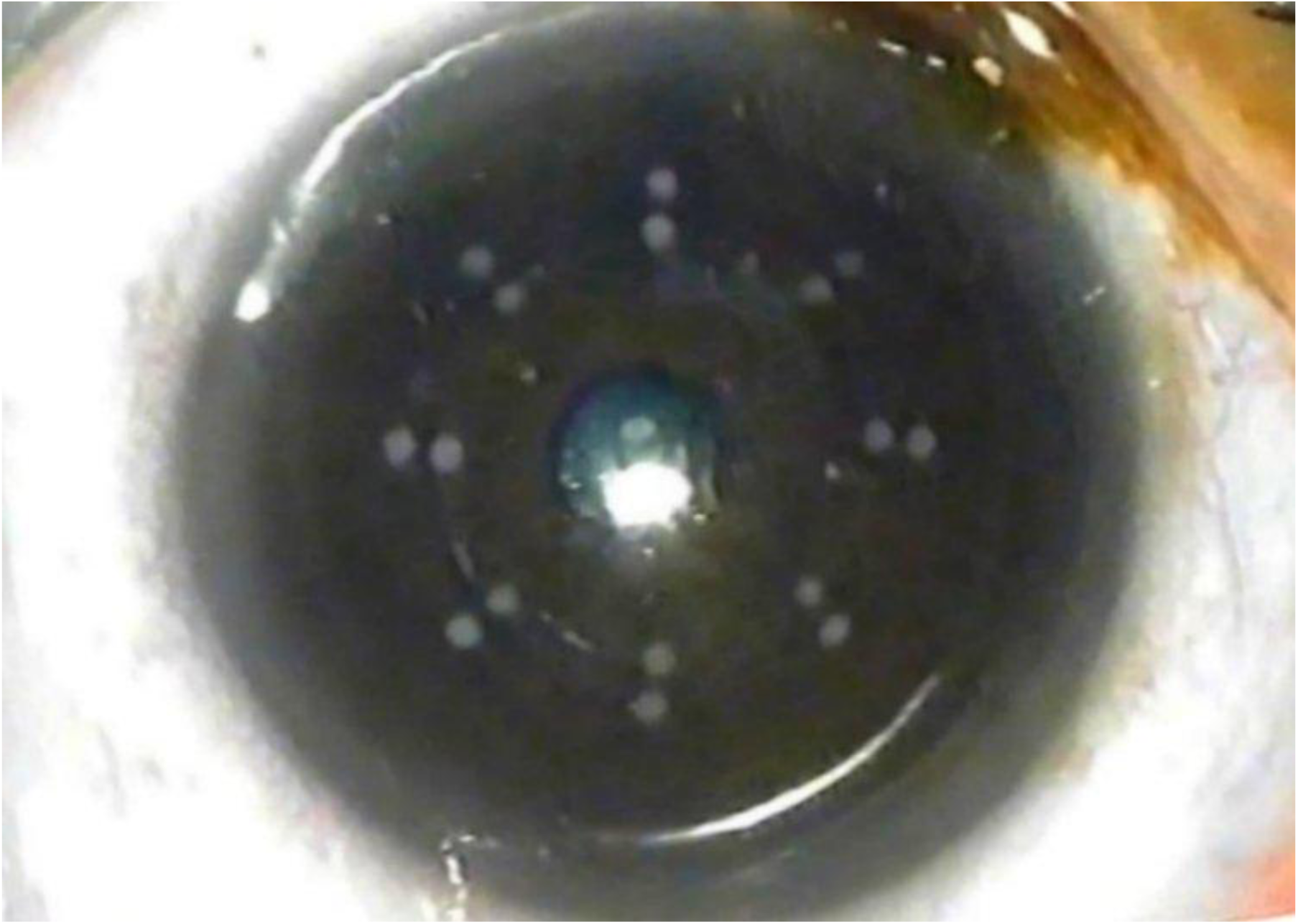
- Eye image immediately post-primary Tx.

The Tx procedure was as follows (see also https://viscorrect.com/wp-content/ uploads/ 2024/09/Opti-K-Procedure.mp4 for an animation):

1. – the supine patient looked upward,
2. – one drop of 0.5% proparacaine HCl was instilled onto the eye to be treated,
3. – a lid speculum was used to hold eyelids open,
4. – a drop of solute-free distilled water was instilled to prevent drying,
5. – the SAWSR was mounted with centration on the pupil center,
6. – the handpiece was docked onto the SAWSR using alignment magnets,
7. – laser energy delivery was completed within 2.5 seconds and
8. – the SAWSR and handpiece were removed.

The procedure evolved over the course of the clinical study in terms of SAWSR designs; pre-Tx instillation fluid - the final choice was solute-free water; Tx nomogram (based on varying the Tx energy density); and post-Tx medication - the final choice was one drop of Xibrom™ (bromfenac ophthalmic solution 0.09%; Ista Pharmaceuticals, Irvine, CA), a nonsteroidal anti-inflammatory drug (NSAID), instilled onto each treated eye immediately post-Tx and at 12 hour periods thereafter for the first 24 hours. No other medications were routinely used.

The first 12 patients were treated differently in their dominant eyes (for distance vision) compared to their nondominant eyes (for near vision) using a monovision protocol. After it was discovered that post-Tx UDVA for the nondominant eyes was maintained or improved, all eyes were treated using a multifocal vision protocol in which both eyes were treated for “best” post-Tx UNVA. Non-primary Txs were typically given when needed for UNVA improvement following regression of primary Tx UNVA values. Most patients received bilateral Txs; eyes of patients not reported herein had pre-Tx hyperopic presbyopia and are part of a separate cohort.

Excel (Microsoft Corporation, Redmond, WA) was used for data entry and general statistical analysis. Mean ± standard deviation values are listed for outcomes. Statistical significance (SS) of outcomes (post-*vs*. pre-primary Tx) was evaluated using Wilcoxon paired sample signed-rank two-sided tests; single (*) and double (**) asterisks indicate SS at the *p* < 0.05 and *p* < 0.01 levels, respectively.

## RESULTS

Post-secondary Tx time is identified by a prime; for example, a single prime as in 12m′ indicates twelve months post-secondary Tx time.

### Safety

There were no clinically significant adverse events or complications in any eye at any post-Tx time. There were only a few temporary (1 day and/or 1 week) cases of a loss of more than 2 lines of corrected distance visual acuity (CDVA) at any post-primary or post-secondary Tx time. The maximum induced astigmatism was 1.25 D and it was temporary. Mean cylinder changes for all eyes at all post-Tx times were in the range of -0.07 to +0.08 D for primary Txs and -0.03 to +0.31 D for secondary Txs.

Tables 1 and 2 list mean intraocular pressure (IOP) and mean central cornea thickness (CCT) respectively for subsets of eyes. There was no statistically significant (SS; *p* < 0.05) change in mean IOP due to either primary or secondary Txs. No eye experienced IOP increase greater than 2 mm Hg; no eye had IOP > 18 mm Hg at any examination time. There was a statistically significant change (*p* = 0.013) in mean CCT due to primary, but not secondary, Txs. No eye experienced CCT change greater than 46 µm. Mean CCT increase (from 520 ± 26 to 536 ± 33 µm) due to primary Txs was not clinically significant.

**Table 1.** – Emmetropic Presbyope mean ± standard deviation intraocular pressure (IOP) measurements: *n* - number of eyes; longest follow-up (f/u) time (either 12m or 24m) used for post-primary Tx and post-secondary Tx measurements.

| **Parameter** | ***n*** | **Pre-primary Tx** | **Post-primary Tx** | **Post-secondary Tx** |
| --- | --- | --- | --- | --- |
| IOP (mm Hg) | 24 | $14.7 \pm 2.3$ | $14.5 \pm 1.8$ ($p = 0.81$) | |
| | 17 | $15.2 \pm 2.6$ | | $14.5 \pm 2.3$ ($p = 0.48$) |

**Table 2.** – Emmetropic Presbyope mean ± standard deviation central corneal thickness (CCT) measurements: *n* - number of eyes; longest follow-up (f/u) time (either 12m or 24m) used for post-primary Tx and post-secondary Tx measurements. *SS (*p* < 0.05).

| Parameter | *n* | Pre-primary Tx | Post-primary Tx | Post-secondary Tx |
| --- | --- | --- | --- | --- |
| CCT ($\mu\text{m}$) | 10 | $520 \pm 26$ | $536 \pm 33$ ($p = 0.013$)* | |
| | 13 | $519 \pm 13$ | | $521 \pm 12$ ($p = 0.610$) |

#### Epithelial Protection

Superficial punctate keratitis (SPK) was observed (by slit-lamp biomicroscopic examination using Na fluorescein staining) in some Tx spots. Epithelial damage was infrequent immediately post-Tx, was more frequent at the 1d post-primary Tx exam and was negligible or absent at the 1w and later f/u exams. Secondary Txs typically had a lower incidence of SPK compared to primary Txs, partly due to reduced Tx energy density. The procedure evolved over the course of the clinical study to increase epithelial protection and reduce discomfort. It was found that irrigating solutions such as preservative-free (PF) Systane^®^ (Alcon Laboratories, Fort Worth, TX) and PF balanced saline solution (BSS) increased the incidence of SPK, so solute-free water (SFW) was selected as the lubricant of choice. As the study progressed, SPK was found to be negligible for Tx energy density of 48 mJ/spot or less.

### Efficacy

#### Monocular uncorrected distance and near visual acuities (UDVA and UNVA)

Table 3 lists mean monocular uncorrected distance and near visual acuities (UDVA and UNVA) in logMAR, decimal and Snellen values, together with standard deviations of logMAR values, numbers of eyes at each time, and an indication of statistical significance (one asterisk * for p = < 0.05, two asterisks ** for p = <0.01) for 145 emmetropic presbyopia (EP) eyes that received primary Opti-K Txs. Figure 3 shows geometric mean (gm) monocular uncorrected visual acuities (VAs) as a function of time from pre-Tx through 730 days (24m) post-primary Tx Figure 3 shows mean VAs graphed in decimal units, with Snellen equivalents shown to the right of the graph. The red line separates “functional” (above line) from partly “dysfunctional” (below line) outcomes, taking 0.5 decimal (20/40 Snellen) as the “functional” value – for UDVA, the value typically required to obtain a driver’s license without eyewear and for UNVA, the value typically required to read “normal” font size (12 pt) text. Pre-Tx and at all post-primary Tx times, 100% of EP patients had UDVA = 20/40 (“functional”) or better.

**Figure 3.**
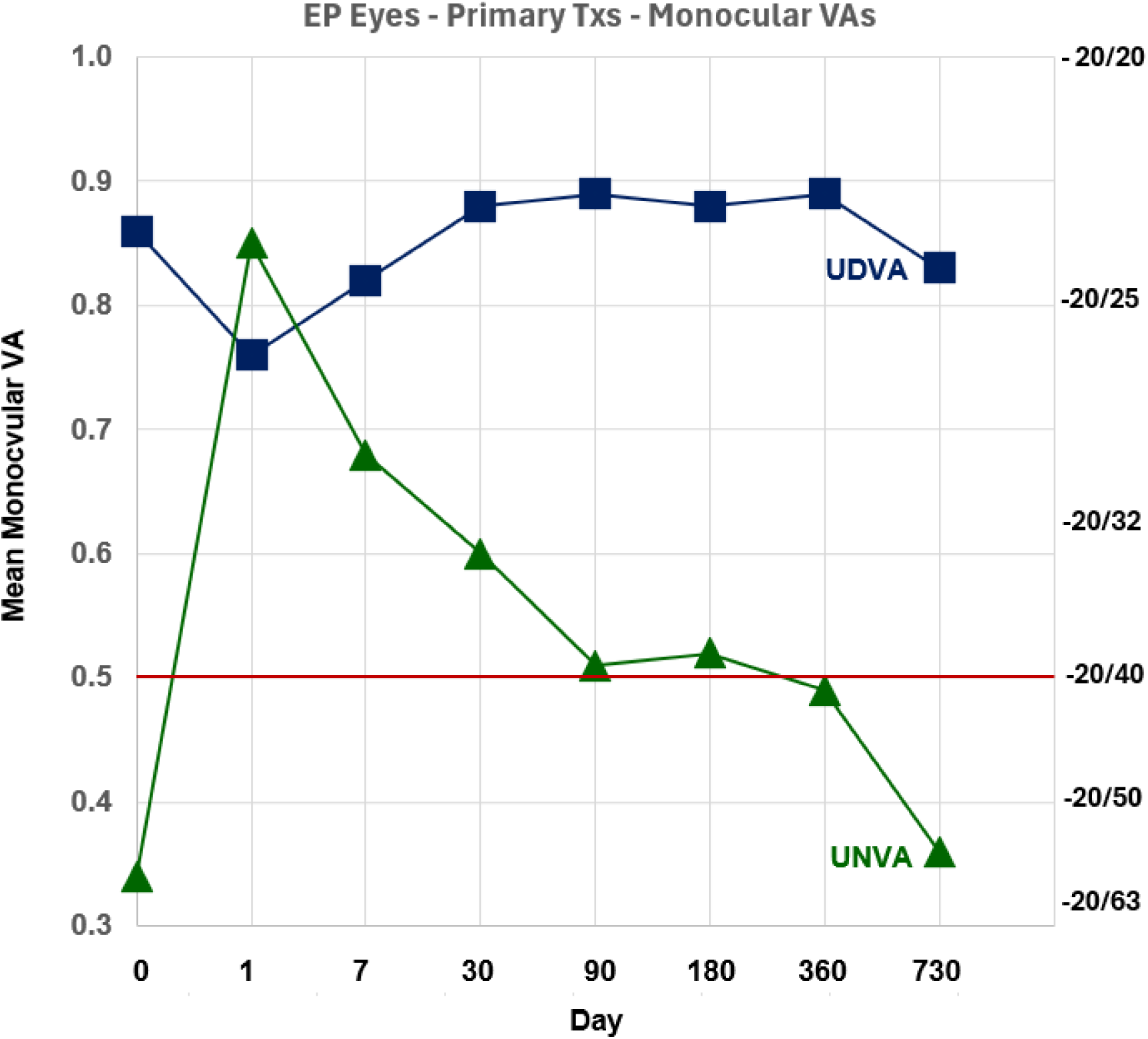
– Geometric Mean Monocular Visual Acuities (decimal value) *vs*. Time for EP eyes that received Primary Opti-K Txs. The red line indicates the “functional” level of VA (20/40). Snellen equivalents are shown on the right of the graph.

**Table 3:** Emmetropic Presbyope Mean (±SD) Monocular Uncorrected Visual Acuities (logMAR values) *vs*. time (upper line) and mean decimal values (middle line) for Primary Opti-K Txs. Snellen values are shown in parentheses.N: number of eyes, Statistical significance: *for p = <0.05, **for p = <0.01.

|  | **Pre** | **1d** | **1w** | **1m** | **3m** | **6m** | **12m** | **24m** |
| --- | --- | --- | --- | --- | --- | --- | --- | --- |
| **UDVA** | 0.07±0.10<br>0.86<br>(20/24) | 0.12±0.18*<br>0.76<br>(20/26) | 0.09±0.13<br>0.82<br>(20/25) | 0.06±0.09<br>0.88<br>(20/23) | 0.05±0.08<br>0.89<br>(20/22) | 0.05±0.08<br>0.88<br>(20/22) | 0.05±0.08<br>0.89<br>(20/22) | 0.08±0.08<br>0.83<br>(20/24) |
| **UNVA** | 0.47±0.20<br>0.34<br>(20/59) | 0.07±0.11**<br>0.85<br>(20/23) | 0.17±0.14**<br>0.68<br>(20/29) | 0.22±0.15**<br>0.60<br>(20/33) | 0.29±0.19**<br>0.51<br>(20/39) | 0.29±0.20**<br>0.52<br>(20/39) | 0.32±0.14**<br>0.48<br>(20/42) | 0.47±0.29<br>0.34<br>(20/59) |
| **N** | 145 | 145 | 127 | 117 | 92 | 60 | 36 | 23 |

Table 4 lists corresponding UDVA and UNVA information in the same format as in Table 3, but for 66 eyes that received both primary and secondary Opti-K Txs. In the same format as Figure 3, Figure 4 shows geometric mean (gm) monocular uncorrected visual acuities (VAs) as a function of time from pre-Tx through 730 days (24m) post-secondary Tx for 66 emmetropic presbyopia (EP) eyes that received both primary and secondary Opti-K Txs. Secondary Txs were typically completed when UNVA decreased below 0.5 (20/40), but some patients received secondary treatments at better than 20/40 UNVA since these patients wanted even better UNVA in order to perform near vision tasks such as reading fine print or threading a needle.

**Figure 4.**
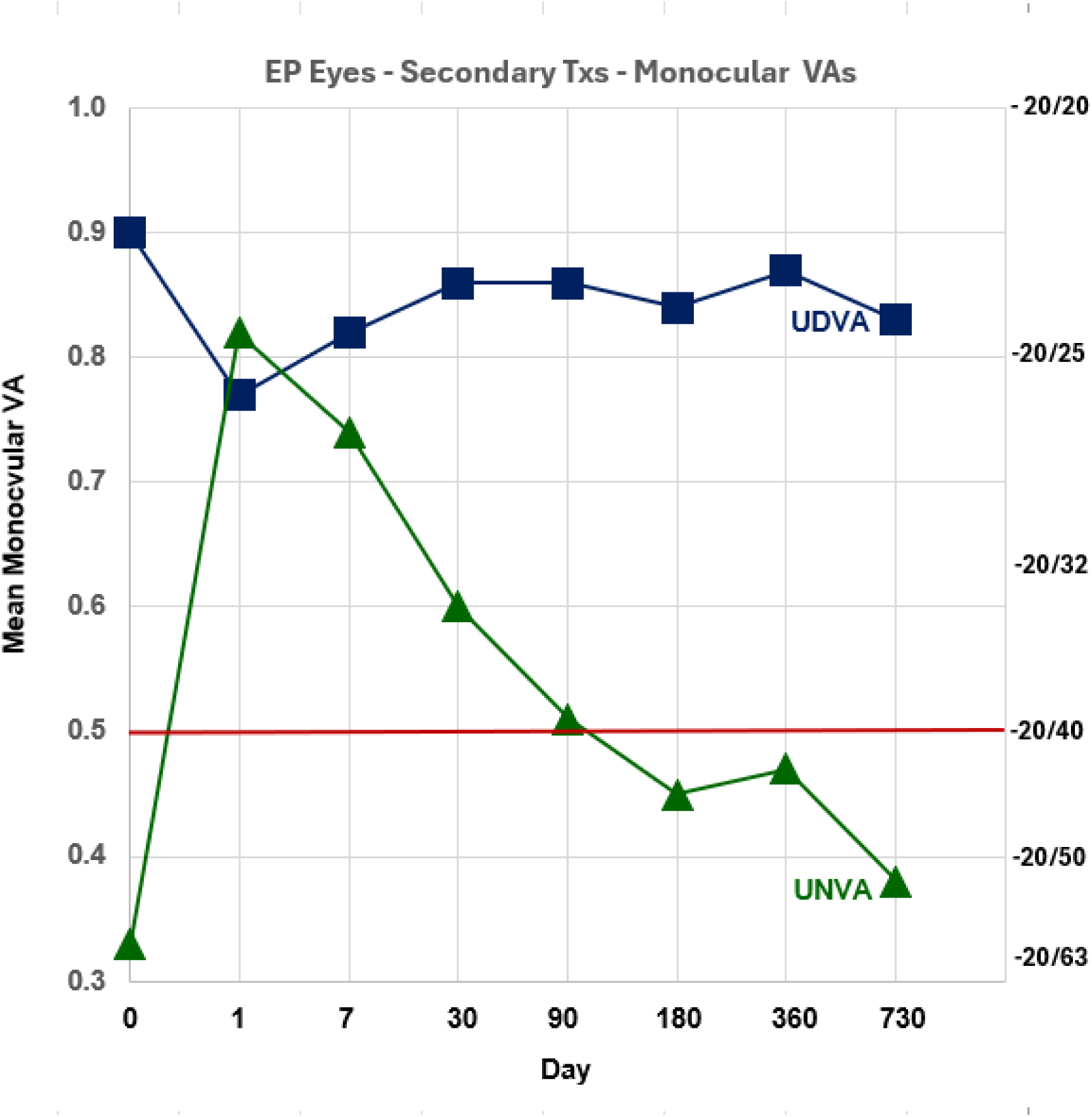
– Same as Figure 3 except for EP eyes that received both primary and Secondary Opti-K Txs.

**Table 4:** Emmetropic Presbyope Mean (±SD) Monocular Uncorrected Visual Acuities (logMAR values) *vs*. time (upper line) and mean decimal values (middle line) for Primary plus Secondary Opti-K Txs. Snellen values are shown in parentheses. N: number of eyes, Statistical significance: *for p = <0.05, **for p = <0.01.

|  | **Pre** | **1d´** | **1w´** | **1m´** | **3m´** | **6m´** | **12m´** | **24m´** |
| --- | --- | --- | --- | --- | --- | --- | --- | --- |
| **UDVA** | 0.05±0.09<br>0.90<br>(20/23) | 0.11±0.23*<br>0.77<br>(20/26) | 0.09±0.14*<br>0.82<br>(20/25) | 0.07±0.09*<br>0.86<br>(20/23) | 0.07±0.08*<br>0.86<br>(20/24) | 0.08±0.07<br>0.84<br>(20/24) | 0.06±0.11<br>0.88<br>(20/23) | 0.08±0.08<br>0.83<br>(20/24) |
| **UNVA** | 0.48±0.19<br>0.33<br>(20/60) | 0.09±0.11**<br>0.82<br>(20/25) | 0.13±0.12**<br>0.74<br>(20/27) | 0.22±0.12**<br>0.60<br>(20/33) | 0.30±0.20**<br>0.51<br>(20/39) | 0.35±0.22<br>0.45<br>(20/47) | 0.32±0.12*<br>0.47<br>(20/42) | 0.42±0.23<br>0.38<br>(20/48) |
| **N** | 66 | 52 | 42 | 33 | 34 | 13 | 25 | 22 |

Too few eyes received ternary or quaternary Opti-K Txs for analysis with statistical significance, but VA trends were similar to primary and secondary Tx cohorts.

#### Binocular uncorrected distance and near visual acuities (Bi UDVA and Bi UNVA)

Table 5 lists mean binocular uncorrected distance and near visual acuities (Bi UDVA and Bi UNVA) in logMAR, decimal and Snellen values, together with standard deviations of logMAR values, numbers of patients at each time, and an indication of statistical significance (* for p = < 0.05, ** for p = <0.01) for 75 emmetropic presbyopia (EP) patients who received primary Opti-K Txs. Figure 5 shows geometric mean (gm) binocular uncorrected visual acuities (Bi VAs) as a function of time from pre-Tx through 730 days (24m) post-primary Tx for 75 emmetropic presbyopia (EP) patients who received primary Opti-K Txs. Figure 5 VAs are graphed in decimal units, with Snellen equivalents shown to the right of the graph. The red line separates “functional” (above line) from partly “dysfunctional” (below line) outcomes, taking 0.5 decimal (20/40 Snellen) as the “functional” value – for Bi UDVA, the value typically required to obtain a driver’s license without eyewear and for Bi UNVA, the value typically required to read “normal” font size (12 pt) text. Pre-Tx and at most post-primary Tx times, 100% of EP patients had Bi UDVA = 20/40 (“functional”) or better. Mean Bi UNVA Snellen lines gained ranged from a peak of 3.2 at 1d post-Tx to 2.1 at 3m post-Tx (the last f/u time for which few secondary Txs were given) to 1.8 at 12m post-Tx (for the remaining patients with primary-only Txs.

**Figure 5.**
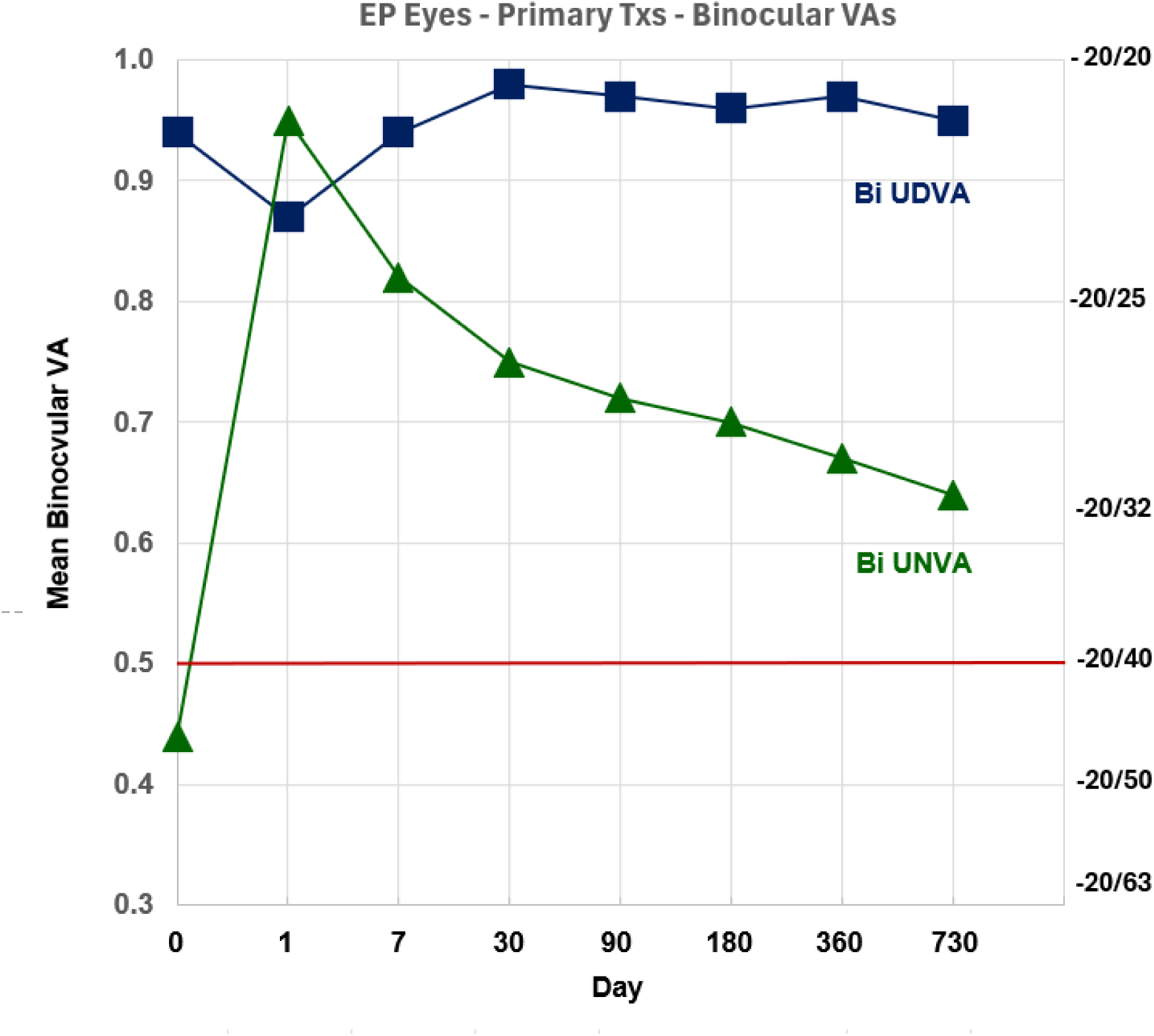
– Geometric Mean Binocular Visual Acuities (decimal values) *vs*. Time for EP patients who received Primary Opti-K Txs. The red line indicates the “functional” level of VA (20/40). Snellen equivalents are shown on the right of the graph.

**Table 5:** Emmetropiic Presbyope Mean (±SD) Binocular Uncorrected Visual Acuities (logMAR values) *vs*. time (upper line) and mean decimal values (middle line) for Primary Opti-K Txs. Snellen values are shown in parentheses. N: number of patients, Statistical significance: * for p = <0.05, ** for p = <0.01.

|  | **Pre** | **1d** | **1w** | **1m** | **3m** | **6m** | **12m** | **24m** |
| --- | --- | --- | --- | --- | --- | --- | --- | --- |
| **Bi UDVA** | 0.03±0.07<br>0.94<br>(20/22) | 0.06±0.12<br>0.88<br>(20/23) | 0.03±0.09<br>0.94<br>(20/22) | 0.01±0.06<br>0.98<br>(20/21) | 0.02±0.06<br>0.97<br>(20/21) | 0.02±0.05<br>0.96<br>(20/21) | 0.01±0.04<br>0.97<br>(20/21) | 0.02±0.04<br>0.95<br>(20/21) |
| **Bi UNVA** | 0.36±0.20<br>0.44<br>(20/46) | 0.02±0.05**<br>0.95<br>(20/21) | 0.09±0.10**<br>0.82<br>(20/24) | 0.13±0.13**<br>0.75<br>(20/27) | 0.14±0.11**<br>0.72<br>(20/28) | 0.15±0.15**<br>0.70<br>(20/29) | 0.17±0.09**<br>0.67<br>(20/30) | 0.20±0.15<br>0.64<br>(20/32) |
| **N** | 75 | 75 | 66 | 63 | 48 | 32 | 25 | 14 |

Table 6 lists corresponding Bi UDVA and Bi UNVA information in the same format as in Table 5, but for 33 EP patients who received both primary and secondary Opti-K Txs.In the same format as in Figure 5, Figure 6 shows geometric mean (gm) binocular uncorrected visual acuities (Bi VAs) as a function of time from pre-Tx through 730 days (24m) post-secondary Tx for 33 emmetropic presbyopia (EP) patients who received both primary and secondary Opti-K Txs. Mean Bi UNVA Snellen lines gained ranged from a peak of 4.0 at 1d post-secondary Tx to 1.8 at 12m post-secondary Tx.

**Figure 6.**
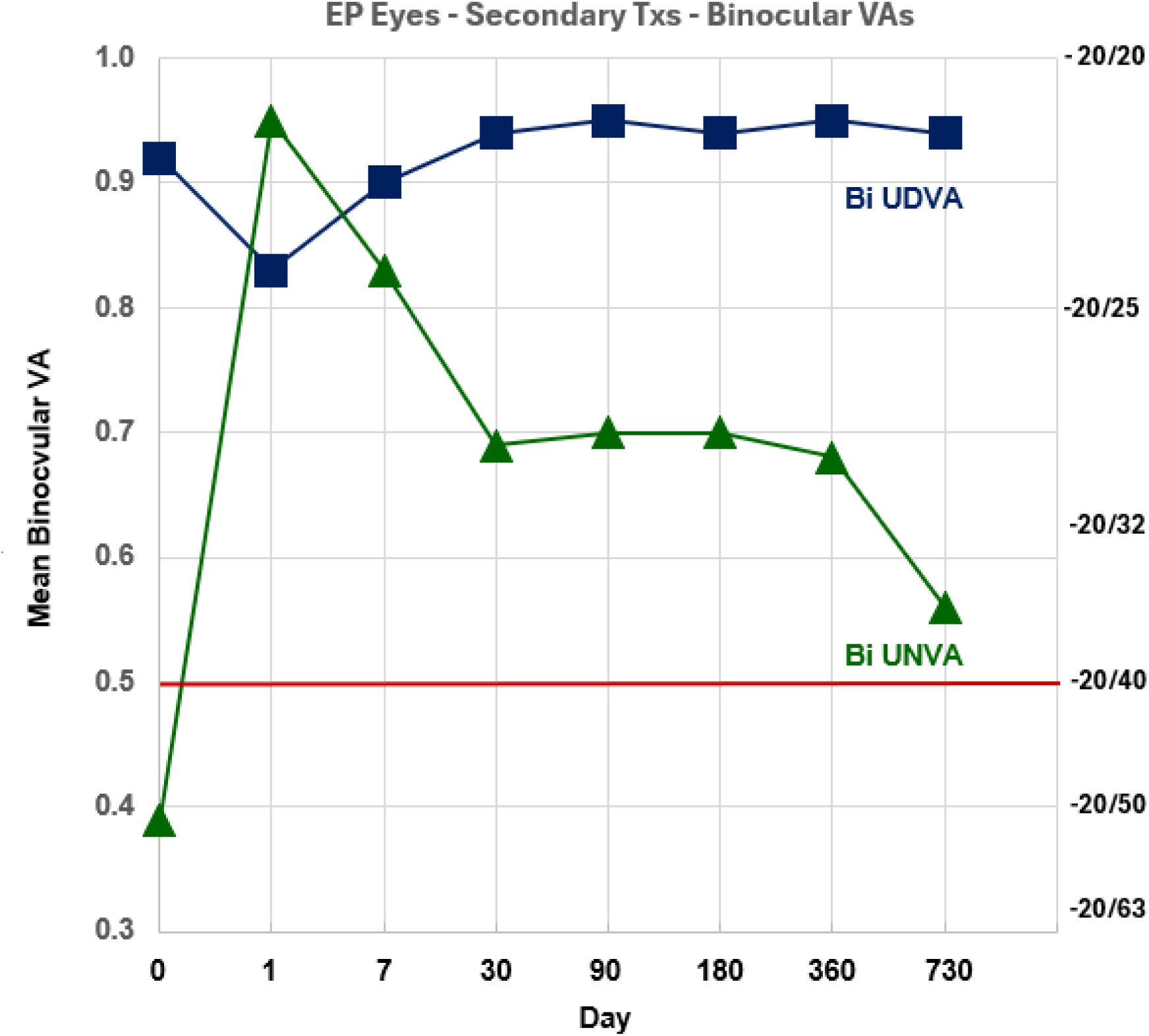
– Same as Figure 5 except for EP patients who received both primary and Secondary Opti-K Txs.

**Table 6.** : Emmetropic Presbyopes Mean (±SD) Binocular Uncorrected Visual Acuities (logMAR values) *vs*. time (upper line) and mean decimal values (middle line) for Primary plus Secondary Opti-K. Snellen values are shown in parentheses. N: number of patients, Statistical significance: * for p = <0.05, ** for p = <0.01.

|  | **Pre** | **1d´** | **1w´** | **1m´** | **3m´** | **6m´** | **12m´** | **24m´** |
| --- | --- | --- | --- | --- | --- | --- | --- | --- |
| **Bi UDVA** | 0.03±0.07<br>0.94<br>(20/22) | 0.08±0.16*<br>0.83<br>(20/24) | 0.05±0.10<br>0.90<br>(20/22) | 0.03±0.07<br>0.94<br>(20/22) | 0.02±0.04<br>0.95<br>(20/21) | 0.03±0.05<br>0.94<br>(20/22) | 0.02±0.05<br>0.95<br>(20/21) | 0.02±0.06<br>0.85<br>(20/21) |
| **Bi UNVA** | 0.41±0.20<br>0.39<br>(20/52) | 0.02±0.04**<br>0.95<br>(20/21) | 0.08±0.10**<br>0.83<br>(20/24) | 0.16±0.15**<br>0.69<br>(20/29) | 0.15±0.13**<br>0.70<br>(20/29) | 0.16±0.07**<br>0.69<br>(20/29) | 0.17±0.09**<br>0.68<br>(20/30) | 0.26±0.17**<br>0.56<br>(20/36) |
| **N** | 33 | 33 | 24 | 20 | 23 | 10 | 14 | 13 |

Too few eyes received ternary or quaternary Opti-K Txs for analysis with statistical significance, but Bi VA trends were similar to primary and secondary Tx cohorts.

Figure 7 shows the correlation between EP Bi UNVA lines gained for 1d post-Tx vs. EP Pre-Tx Bi UNVA. The linear regression coefficient of determination R² is large (0.80) indicating that the largest lines gained are obtained for the patients with the poorest pre-Tx Bi UNVA. Conversely, only small lines gained are obtained for patients with the best pre-Tx Bi UNVA. Similar correlations were observed at later f/u times.

**Figure 7.**
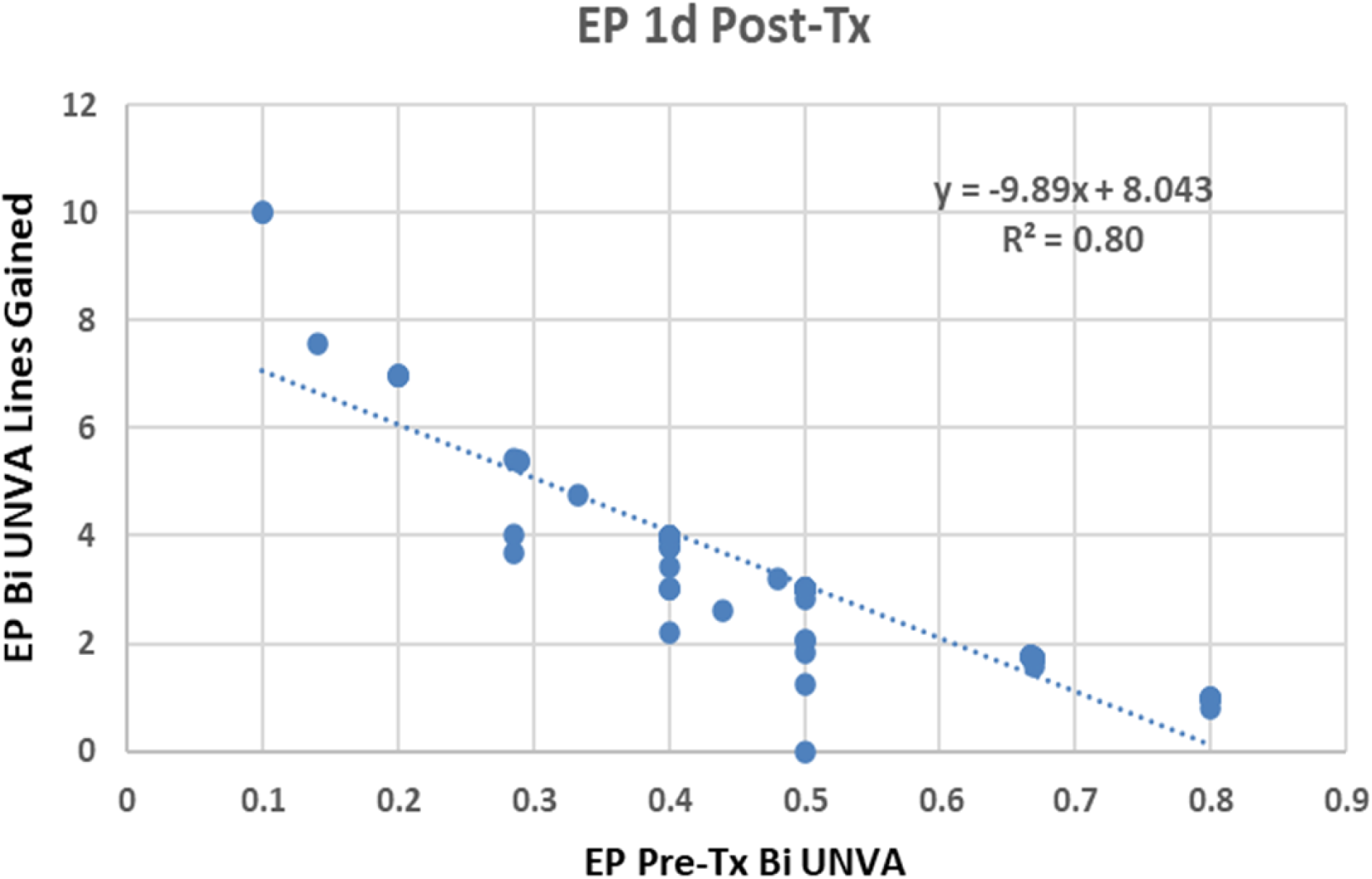
– EP Bi UNVA lines gained vs. EP pre-Tx Bi UNVA (in decimal units), Linear regression fitting coefficients (slope and intercept) and R value are shown in the upper right portion of the figure.

#### Manifest Refraction, Spherical Equivalent (Delta MRSE)

Mean ± SD manifest refraction, spherical equivalent (MRSE) for 145 eyes was +0.28 ± 0.27 D pre-Tx and -0.25 ± 0.48 D at 1 week post-primary Opti-K Tx, corresponding to a MRSE change (myopic shift) of -0.52 ± 0.55 D, and then gradually returned nearly to the pre-primary Tx value at 24m post-Tx. Figure 8 shows changes (relative to pre-Tx) for mean MRSE values as a function of time from pre-Tx through 730 days (24m) post-primary Tx for 145 emmetropic presbyopia (EP) eyes that received primary Opti-K Txs. Changes (relative to pre-Tx) for mean Delta MRSE values for EP eyes that received both primary and secondary Opti-K Txs have a similar time course as in Figure 7 but with a minimum value of -0.31 D at 1 week post-Tx.

**Figure 8.**
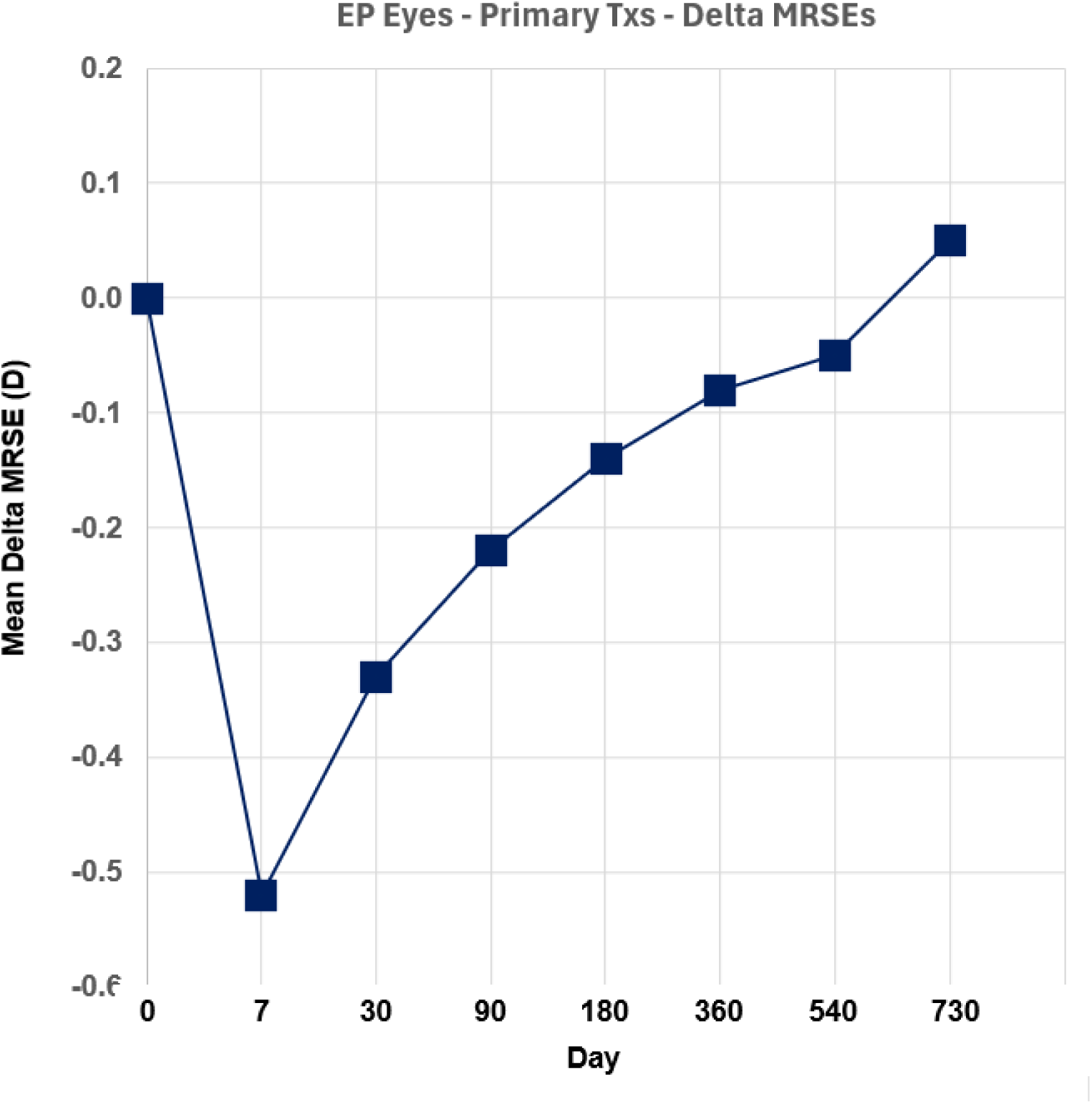
– Mean Change in Manifest Refraction, Spherical Equivalent (Delta MRSE) *vs*. time for EP eyes that received primary Opti-K Txs.

#### Corneal Topography (CT)

Corneal topography (CT) maps showed mean ± SD central steepening of 0.51 ± 0.49 D from pre-Tx to 1 week post-primary Tx, in good agreement with the mean ± SD MRSE change (myopic shift) of -0.52 ± 0.55 D. At 12m post-primary Tx, the CT steepening was 0.27 ± 0.38 D, somewhat larger than the MRSE change of -0.08 ± 0.33 D. Figure 9 shows an example of the unusual CT difference (post-*vs*. pre-Tx) map that was observed in some eyes at early f/u times. The CT difference map shows alternating steeper and flatter radial segments that radiate from the Tx pattern center to the paracentral zone (*ca*. 6 mm diameter) where the inner ring of Tx spots is located (*cf*. Figure 2). The pattern of corneal shape change is approximately that of an eight-leaf rosette, with bisected leaves having steeper and flatter sectors. This multifocal corneal shape is the basis of multifocal vision in eyes treated by Optimal Keratoplasty (Opti-K).

**Figure 9.**
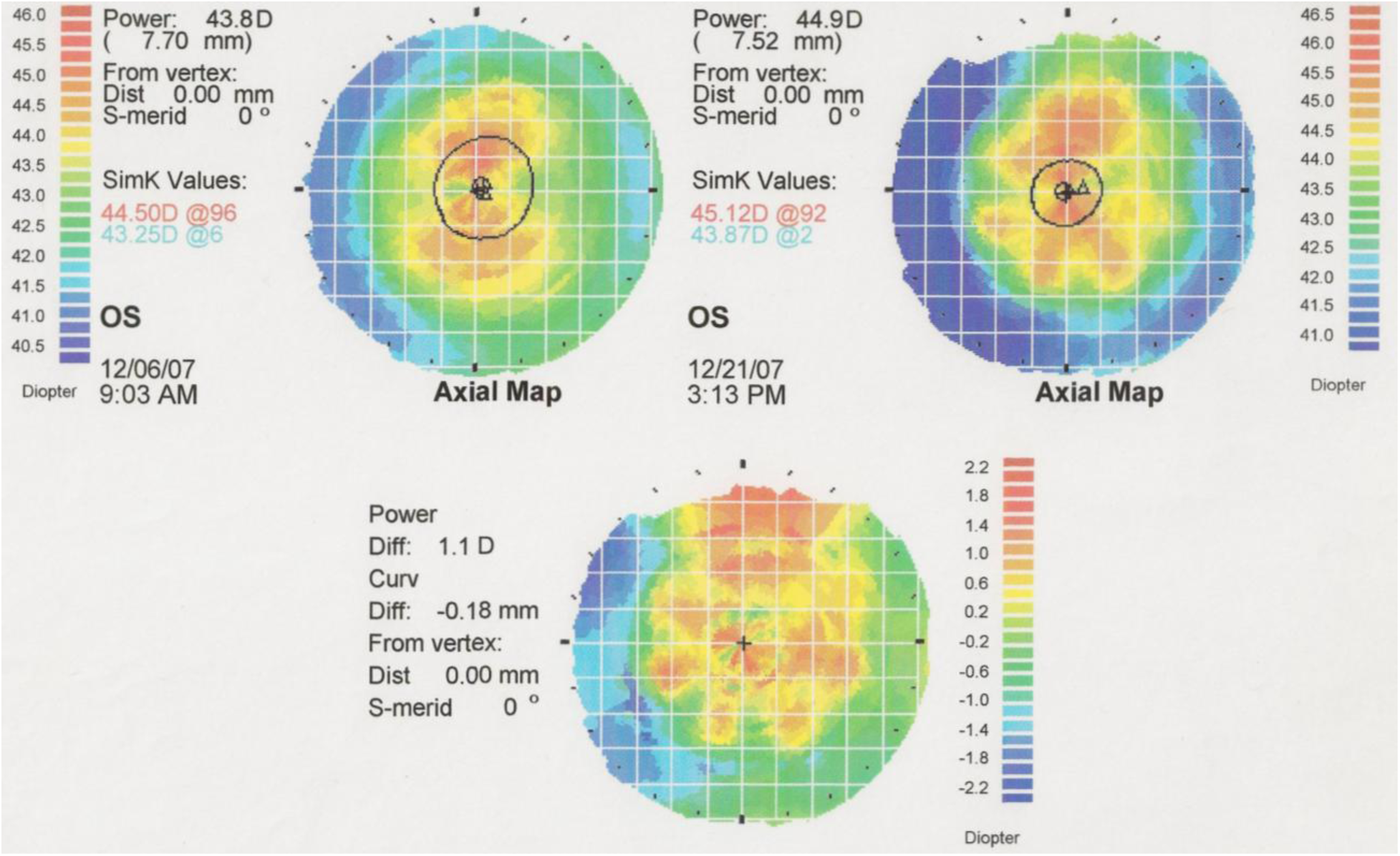
– Corneal topography maps. Top left: pre-primary Tx. Top right: 1w post-primary Tx. Bottom: difference (1w post-minus pre-Tx).

#### Comfort

Subjective patient self-assessments of discomfort due to pain, tearing, photophobia and foreign body sensation (fbs) were each obtained on a scale of 0 (none), 1 (trace), 2 (mild), 3 (moderate) and 4 (severe). Maximum discomfort occurred on 1d post-primary Tx, with means ± standard deviations of 0.22 ± 0.57 (pain), 0.31 ± 0.83 (tearing), 0.57 ± 1.11 (photophobia) and 0.65 ± 1.11 (fbs) for all eyes. Patients who received Xibrom™ immediately post-Tx and at 12 hour intervals for 24 hours post-Tx had little or no discomfort. Other procedure improvements may also have contributed to comfort increase - the last 25 EP eyes that received primary Txs had 1 eye with 1 (pain), 0 (tearing), 2 eyes with 1 (photophobia) and 7 with 1 and 1 with 2 (fbs) on 1d post-primary Tx. Comfort was typically better for secondary Txs compared to primary Txs due, in part, to reduced Tx energy density. Comfort was usually excellent at 1w, 1w′ and later post-Tx times.

#### Recovery Time

Most patients had little or no post-Tx recovery time. As evidenced by visual acuity and subjective comfort outcomes, the majority of patients were fully functional at 1 day post-Tx.

#### Patient Satisfaction with Binocular Distance and Near Vision

Subjective self-assessments of patient satisfaction with binocular distance vision (SDV) and binocular near vision (SNV) were made on a 5-point scale in which a rating of 5 corresponded to high satisfaction and other ratings were 4 - satisfaction, 3 - neither satisfaction nor dissatisfaction, 2 - dissatisfaction, and 1 - high dissatisfaction. Baseline (pre-primary Tx) mean SDV was high: 4.30 ± 0.73 while mean SNV was low: 2.08 ± 0.76.

For mean SDV: Post-primary Tx values indicated greater satisfaction than at baseline at most f/u times. Post-secondary Tx values indicated less satisfaction at 1w′ (due to some overcorrection cases), but greater satisfaction at longer f/u times. None of the mean SDV changes were statistically significant (*p* < 0.05).

For mean SNV: Post-primary Tx values indicated much greater satisfaction than at baseline; values at 1d (4.45 ± 0.68; \**p<* 0.01), 1w (3.90 ± 0.93; \*\**p* < 0.01), 1m (3.64 ± 0.98; \*\**p* < 0.01) and 6m (3.30 ± 0.95;\**p* < 0.05) had statistically significant (SS) improvements relative to baseline. Post-secondary Tx values also indicated greater satisfaction than at baseline; values at 1d′ (4.46 ± 0.74;\**p*<0.05), 1w′ (3.78 ± 0.73; \**p*<0.05), 1m′ (3.75±0.63;*p<0.05), and 3m′ (3.14 ± 0.76; \**p<*0.05) had SS improvements but at later f/u times, mean SNV values declined further and were not SS, declining to 24m′ (2.50±1.14). Other post-primary and post-secondary Tx value improvements in mean SNV did not have SS, partly due to small numbers of patient responses at those f/u times.

## DISCUSSION

### Visual Acuities

Distance VA (both UDVA and Bi UDVA; for both primary and secondary Txs) was conserved, with few statistically significant temporary changes from baseline. Near VA (both UNVA and Bi UNVA; for both primary and secondary Txs) improvements were large and very statistically significant (mostly p << 0.01) at early f/u times but regressed so that additional Txs were typically needed to boost improvement. Bi UNVA outcomes were considerably better than UNVA outcomes, due to binocular interaction [Ref. 8]. The mean ± SD duration of “functional” Bi UNVA effect (with mean value 20/40 or better) was 10.8 ± 9.1 months for patients with primary Txs only but 33 of 75 (44.0%) patients required secondary Txs. The mean ± SD duration of “functional” Bi UNVA effect was 15.2 ± 10.3 months for patients with both primary and secondary Txs. Regression of duration of effect timescales were confirmed by MRSE and CT measurements; both showed that initial refractive change and corneal steepening decreased as f/u time increased. There were no statistically significant correlations of Bi UNVA outcomes with patient gender or age; however, there was a significant correlation of outcomes (in terms of Bi UNVA lines gained) *vs*. pre-Tx Bi UNVA (Figure 7).

Presbyopia is a progressive condition, typically requiring presbyopic patients to change eyewear every 2 to 3 years to improve near vision. The duration of Opti-K treatment (Tx) effect is less than needed to match the full timescale of presbyopia progression; however, the procedure is so safe, noninvasive, comfortable and repeatable that Opti-K retreatment is a fine option. In addition, in contrast to many other procedures for presbyopia correction that involve monovision [Ref. 3], Opti-K Txs are typically given bilaterally, thereby retaining stereoacuity, contrast sensitivity and freedom from dysphotopsias such as glare and halo.

Opti-K visual acuity outcomes presented herein are consistent with previous Opti-K clinical study outcomes [Refs. 6 and 7]. Outcomes for a different thermal procedure for near vision improvement for emmetropic presbyopes (EPs), conductive keratoplasty (CK), have been reported [Refs. 9 and 10]. The CK outcomes are not directly comparable to the present Opti-K outcomes since CK Txs were all unilateral in a monovision protocol. Monovision was adopted for EPs probably due to the high percentages of CK-treated eyes that lost significant uncorrected distance visual acuity – 77% and 60% had UDVA of 20/25 or worse at 3 months and 1 year post-Tx, respectively; 33% had UDVA of 20/50 or worse (dysfunctional) at 3 months post-Tx [Ref. 10 – Figure 28]. As emphasized herein, Opti-K Tx conserves UDVA.

### Study Limitations

The present Opti-K clinical study is limited in terms of the number of patients (75), follow-up time (2 years or less), and lack of other measurements (specular microscopy, reading speed, contrast sensitivity, stereoacuity, aberrometry, histology, *etc*.) that would contribute to understanding the mechanism of action and full outcomes of Optimal Keratoplasty treatment.

### Conclusions

Optimal Keratoplasty (Opti-K) appears to be safe and efficacious in providing temporary improvement of near visual acuity to patients with emmetropic presbyopia while maintaining (or improving) their distance visual acuity by a multifocal vision protocol. The Opti-K Tx procedure is completely noninvasive and rapid and is simple and convenient for both physician and patient. The procedure has evolved to incorporate secondary Txs as needed to further improve near vision. Bi UNVA improvement to a functional decimal level of 0.50 (0.30 logMAR, 20/40 Snellen) has a duration of more than two years post-secondary Tx for at least half the patients, which matches the “temporary prescription” needed by many patients in the presbyopic age range.

Although temporary, Opti-K treatments are repeatable and can be applied whenever needed to maintain both functional near and distance visual acuities. The unique Opti-K treatment pattern has eight-fold symmetry that provides optimal multifocal vision with immediate neuroadaptation.

## ACKNOWLEDGMENTS

The authors gratefully acknowledge the contributions of Philip Bassett, OD, Yvonne Rolle, Patricia Rodgers, Richard Menefee, Jerry Rein, John Werra and Robert Werra toward the completion of this study.

## DATA AVAILABILITY

The data that support the findings of this study are available on request from the corresponding author. The data are not publicly available due to privacy or ethical restrictions.

## REFERENCES

1 – Holden BA, Fricke TR, Ho SM, et al. Global vision impairment due to uncorrected presbyopia. Arch Ophthalmol. 2008; 126:1731–1739.

2 – Pallikaris IO, Plainis S, Charman WN. Hyperopia – origins, effects and treatment. Slack Inc., Thorofare NJ, 2013.

3 – McDonald MB, Mychajlyszyn A, Mychajlyszyn D, Klyce SD. Advances in corneal surgical and pharmacological approaches to the treatment of presbyopia. J Refract Surg 2021;3716 SupplI:S20–S27.

4 – Klyce SD, McDonald MB. A new approach to thermal keratoplasty. Cataract Refract Surg Today Jan 2024:50-52, 64.

5 – NTK Enterprises, Inc. (now VIS, Inc.). Clinical study report: feasibility study of presbyopia correction using the NTK optimal keratoplasty (Opti-K™) system. IDE G170310 – 2021 report.

6 – Rodgers KJ, Glen HG, Salz JJ, et al. Improved method of laser thermal keratoplasty to overcome presbyopia. In: Ophthalmic Technologies XXI, edited by Manns F, Söderberg PG, Ho A. Proc SPIE 2011;7885:N-1 through N-8.

7 – Veritti D, Sarao V, Lanzetta P. Optimal keratoplasty for the correction of presbyopia and hypermetropia. J Ophthalmol 2017;ID 7545687:1–6.

8 – Yassin M, Lev M, Polat U. Space, Time and Dynamics of Binocular Interactions. Sci Repts 2023;13:21449. Available at https://www.nature.com/scientificreports.

9 – McDonald MB, Durrie D, Asbell P, et al. Treatment of presbyopia with conductive keratoplasty – six-month results of the 1-year United States FDA clinical trial. Cornea 2004;23:661–668.

10 – Hersh PS. Optics of conductive keratoplasty: implications for presbyopia management. Trans Am Ophthalmol Soc 2005;103:412–456.

